# A Genomic Analysis of Usher Syndrome: Population-Scale Prevalence and Therapeutic Targets

**DOI:** 10.1101/2025.02.27.25323008

**Authors:** Shelby E. Redfield, Stephanie A. Mauriac, Gwenaëlle S. Géléoc, A. Eliot Shearer

**Affiliations:** Department of Otolaryngology and Communication Enhancement, Boston Children’s Hospital, Boston, MA, USA; Children’s Rare Disease Collaborative, Department of Information Technology, Boston Children’s Hospital, Boston, MA, USA

**Keywords:** Hearing loss, deafness, retinitis pigmentosa, deaf-blindness

## Abstract

Usher syndrome (USH), the most common form of deaf-blindness, displays extensive genetic, allelic, and phenotypic heterogeneity. The dual sensory impairment associated with this disorder makes Usher syndrome an important target for gene therapy, with dozens of published preclinical studies targeting multiple USH genes and using multiple gene therapy strategies. Nine genes have been conclusively linked to Usher syndrome; however, data on prevalence and contribution of specific genetic variants is lacking. Such information is essential to choosing a favorable target gene or therapeutic approach during clinical trial design. Here, we used large genomic databases to systematically evaluate the genomics of Usher syndrome. We ascertained pathogenic USH variants from three clinical databases and determined the occurrence of these pathogenic USH variants within: (1) a publicly available dataset including worldwide populations (GnomAD), (2) a cohort of 3,888 children without hearing loss, and (3) 637 children with hearing loss. Results show significant variability in the frequency of USH variants by gene and genetic ancestry. 1% of control subjects carry a pathogenic USH variant. Pathogenic variants in *USH2A* are the most prevalent, at 1 in 150 individuals (0.0062). Calculated general population prevalence for all USH subtypes is 1 in ∼29,000, indicating that 30,405 individuals in the United States and 721,769 individuals worldwide are affected. We estimate that 324 babies in the United States and 12,090 worldwide are born with Usher syndrome each year. We identify key targets for genetic therapy based on population-level prevalence including a focus on alternatives to gene replacement therapies, specifically for *USH2A*.

## INTRODUCTION

Usher syndrome is the most common syndromic form of hearing loss and the most common cause of hereditary deaf-blindness. Hearing and vision loss are due to progressive degeneration and subsequent loss of cochlear hair cells and retinitis pigmentosa, respectively. Individuals with Usher syndrome are typically born with hearing loss and experience a progressive loss of peripheral vision in childhood or early adulthood. There is notable genetic and phenotypic heterogeneity in Usher syndrome: USH1, the most severe form, is caused by pathogenic genetic variants in *MYO7A, USH1C, CDH23, PCDH15,* and *USH1G*; USH2, associated with later onset vision loss and less severe hearing loss, is caused by pathogenic variants in *USH2A, ADGRV1,* and *WHRN*; while USH3, associated with a more variable phenotype, is caused by pathogenic variants in *CLRN1*. There are several other proposed Usher syndrome genes, but none to date have been unequivocally associated with disease^1^.

The dual sensory impairment of Usher syndrome can have devastating effects on individuals’ ability to communicate, interact socially, and interact with the environment. Hearing aids and cochlear implants can provide access to sound, but do not restore natural hearing. Reduction of UV exposure and Vitamin A supplementation may delay onset but do not prevent retinitis pigmentosa. Given the lack of curative treatment, Usher syndrome is a key focus of new gene and molecular therapies^2,3^.

However, development of these new therapies has been hampered by a lack of accurate prevalence estimates of Usher syndrome. Moreover, there is a lack of knowledge as to how much any individual pathogenic variant contributes to overall USH disease burden. Prior prevalence estimates of 1 in 6,000 to 1 in 16,000 individuals are based on case reports and small groups (< 50) of affected individuals^4–6^. A meta-analysis of 2,476 individuals with sensorineural hearing loss calculated that Usher syndrome was diagnosed in 7.5%^7^. To date, there have been no large studies estimating the prevalence of Usher syndrome variants in control populations. Lack of accurate data for both overall prevalence of Usher syndrome and relative contribution of individual genetic variants precludes impactful, focused development of genetic therapies that prioritizes the appropriate therapy modality.

While adeno-associated viral (AAV) gene replacement therapy holds great promise in the treatment of rare disease and can be applied across any number of individual causative variants in a gene, gene replacement therapy using this viral vector can only be explored for genes that have a coding sequence below 4.8 kb. Alternative approaches must be considered for larger genes. Dual AAV strategies have been successfully developed that take advantage of splitting approaches either at the DNA or protein level^8^. However, several USH genes encode cDNA that extend even beyond dual AAV capacity including *CDH23* (10,062bp), *USH2A* (15,606bp), and *ADGRV1* (18,918bp). As such, alternative therapies such gene editing or use of small molecules or antisense oligonucleotides (ASOs) must be explored to target mutations in any of these three genes.

A primary weakness of therapies that target specific mutations, rather than whole genes, is that the population that stands to benefit is reduced. Prioritization of variants for targeted therapy will depend on several factors including the genetic variant pathogenicity and targetability, the onset and progression of the disease, and the prevalence of said pathogenic variant. USH2 is known to be the most common form; however USH1 typically demonstrates a more severe phenotype. The prevalence of Usher syndrome and specific pathogenic variants that could be targeted, is, to date, not clearly delineated.

The goals of this current study are to (1) systematically assess the carrier frequency and prevalence of Usher syndrome using analysis of large genomic databases in multiple populations worldwide, and (2) assess the frequency of variants in Usher syndrome genes appropriate for targeted genetic and molecular therapies.

To do so, we ascertained pathogenic Usher syndrome variants from three different clinical databases and determined the occurrence of these pathogenic Usher variants within three large genomic databases: (1) a publicly available dataset including worldwide populations (GnomAD), (2) a cohort of 3,888 children without documented hearing loss who underwent exome sequencing (ES) for other medical conditions, and (3) 637 children with hearing loss who underwent targeted gene panel test, ES, and/or GS.

We find significant variability in genetic contribution to Usher syndrome by race and ethnicity. We determine prevalence of all types of Usher syndrome in the United States and worldwide using five different methods for statistical estimation. Finally, we characterize mutational burden of Usher syndrome by type and prevalence and propose therapeutic mechanism. We hereby present the most accurate estimate of Usher syndrome prevalence to date, and provide data that should guide future therapeutic focus.

## METHODS

### Editorial Policies and Ethical Considerations

All human subjects related work in this study was performed with approval from the Boston Children’s Hospital Institutional Review Board and in accordance with their policies. All participants or their parent/guardian provided written informed consent.

### Genetic Variants

Pathogenic and likely pathogenic (P/LP) variants in the genes *MYO7A, USH1C, CDH23, PCDH15, USH1G, USH2A, ADGRV1, WHRN,* and *CLRN1* were ascertained from three databases: (1) ClinVar, 1-3 star annotated only, (2) “Disease Causing Variants”, from Human Gene Mutation Database (HGMD) and (3) the Deafness Variation Database^9^. Variants with conflicting interpretations of pathogenicity were not included.

We assessed both (1) all P/LP variants within Usher syndrome genes as well as (2) only variants with annotated with an Usher syndrome phenotype. These variants were annotated using ANNOVAR^10^ and VEP^11^ and minor allele frequencies were gathered from the April 2024 version of gnomAD (4.1) which includes genomic data from 807,162 individuals (730,947 exomes and 76,215 genomes) from 8 racial/ethnic populations (AFR, African/African American; AMR, Admixed American; ASJ, Ashkenazi Jewish; EAS, East Asian; FIN, Finnish; MID, Middle Eastern; NFE, Non-Finnish European; SAS, South Asian)^12^. Of note, the population ‘Remaining’ was not included in our analysis. For exonic/splice site variants, gnomAD ES was used, whereas gnomAD GS was used for intronic variants given higher coverage levels.

### Control Population

We ascertained genomic data from a large population of children without hearing loss as part of a precision medicine initiative called Children’s Rare Disease Cohorts (CRDC), which has previously been described^13,14^. The CRDC leverages ES and GS data from cohorts of pediatric patients with rare diseases. A genomic learning system (GLS) integrates clinical phenotypic data by combing the electronic health record (EHR) and developing pertinent human phenotype ontology (HPO) terms^13^. For this study, we evaluated only unrelated probands (no family members) and excluded patients with HPO terms related to any form of hearing loss using the “hearing impairment” term (HP:0000364) and all of its descendants in the ontology tree. These individuals had ES performed in a CLIA certified environment. Analysis of genomic data was performed using the DRAGEN pipeline (Illumina Inc., San Diego, CA). Variant analysis and filtering was performed using SeqMiner (GeneDx Inc., Gaithersburg, MD) and variants were annotated with ANNOVAR.

### Affected Population

As part of an ongoing study of pediatric hearing loss, we have ascertained a large cohort of children (637 to date) with likely genetic sensorineural hearing loss. These children underwent targeted gene panel, ES, and/or GS in a CLIA certified environment. As previously reported, the diagnostic yield is 40.7% for bilateral sensorineural hearing loss^15^. This study is approved by the Boston Children’s Hospital IRB and informed consent was obtained.

### Calculation of Prevalence

We calculated prevalence of Usher syndrome (including each Usher subtype) using five different databases of P/LP variants:

1. ClinVar_USH: P/LP variants in all Usher genes from ClinVar, restricted to only variants identified as causing Usher syndrome, annotated with minor allele frequency (MAFs) from gnomAD.
2. ClinVar_All: P/LP variants in all Usher genes from ClinVar, not specifying the phenotype, annotated with MAFs from gnomAD.
3. DVD_All: P/LP variants in all Usher genes from the Deafness Variation Database v9^9^, not specifying the phenotype, annotated with MAFs from gnomAD. Variants with conflicting interpretation from ClinVar (i.e. those classified as Benign/Likely Benign) were excluded from further analyses.
4. HGMD_All: P/LP variants in all Usher genes from HGMD^16^, a commercial database of pathogenic human variation, not specifying the phenotype, annotated with MAFs from gnomAD. As above, variants with conflicting variant interpretation in ClinVar were excluded.
5. gnomAD_LoF: All LoF variants within Usher syndrome genes were identified. We also obtained all loss of function (LoF) variants from gnomAD to provide another estimate of P/LP variant burden within Usher genes. Given that these were candidate variants (i.e. not all were categorized as pathogenic), LoF variants with minor allele frequency (MAF) above the highest MAF of the previously categorized P/LP within that gene were excluded from further analysis. As above, variants with conflicting variant interpretation in ClinVar were excluded.

Prevalence was calculated using the Hardy-Weinberg equation and the sum total of MAFs per gene, then summed within each Usher subtype. We did not account for digenic inheritance of Usher syndrome in these calculations. United States population, world population, United States birth rate, and world birth rate were estimated as 337,000,000, 8,000,000,000, 3,596,017 per year, and 134,000,000 per year, respectively (census.gov and cdc.gov).

Finally, we also calculated adjusted Usher syndrome prevalence using previously derived equation^4^: *y* = (1-*c*) * (*x*/*s*) where *x* = observed frequency, *s* = sensitivity of assay, and *c =* carrier rate. Carrier rate (*c*) was calculated as 0.01, the summed MAF of the most conservative variant database, ClinVar_USH. Sensitivity (*s)* of current Usher syndrome genetic testing assays was determined to be 0.912^7^. Observed frequency of affected individuals was set as 0.065, a combination of data from our affected hearing loss population (637), and 2,476 individuals with hearing loss from Jouret et al^7^.

## RESULTS

### Usher syndrome pathogenic variant allele frequency

We first assessed the prevalence of P/LP variants within Usher syndrome genes within the gnomAD database (**Figure 1**). Comparison of P/LP variant MAFs within Usher genes and between populations with different genetic ancestry demonstrated notable differences. For instance, P/LP variants in *USH1C* and *WHRN* were relatively common in NFE, but essentially non-existent in ASJ, EAS, FIN, and MID populations. P/LP variants in *PCDH15* and *ADGRV1* were common in the ASJ population and not in others, whereas P/LP variants were more evenly distributed across populations for *CDH23* and *USH2A*.

**Figure 1.**
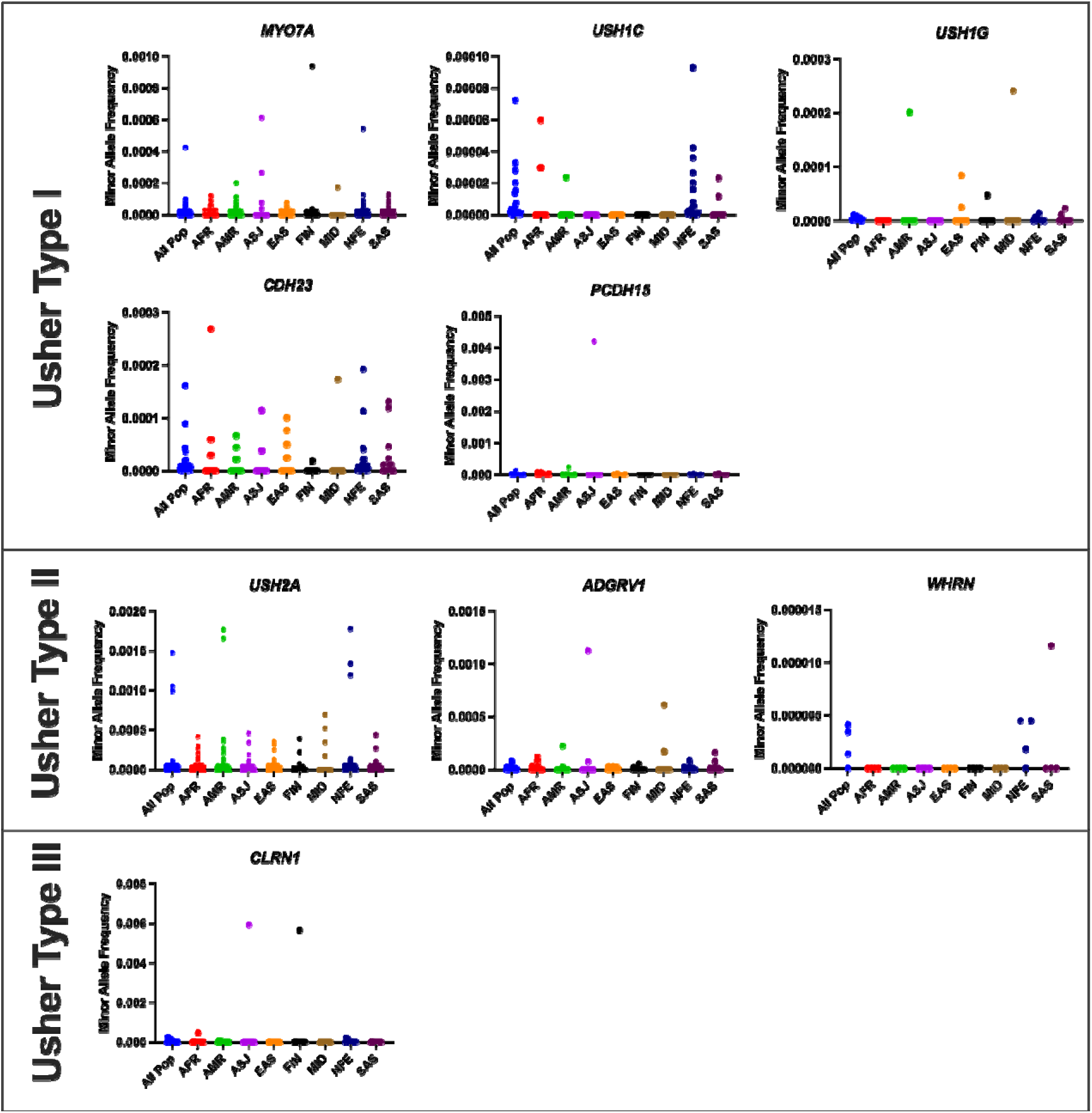
Pathogenic and likely pathogenic Usher syndrome variants allele frequency distribution differs by genetic ancestry. Each gene is shown with populations along the x-axis and minor allele frequency on the y-axis. All pop, all populations; AFR, African/African American; AMR, Admixed American; ASJ, Ashkenazi Jewish; EAS, East Asian; FIN, Finnish; MID, Middle Eastern; NFE, Non-Finnish European; SAS, South Asian. Note, y axes are different scale.

Overall, P/LP variants in *USH2A* were the most common when all populations were combined, followed by *MYO7A* (**Figure 2**). In fact, the top three most common P/LP Usher variants are *USH2A* variants (**Table 1**). *USH2A* also contained 45% of all reported Usher syndrome P/LP variants (575 of 1,276 variants, **Table 2**). While P/LP variants in *PCDH15* were relatively common within the ASJ population, this gene’s overall contributions in all populations were low.

**Figure 2.**
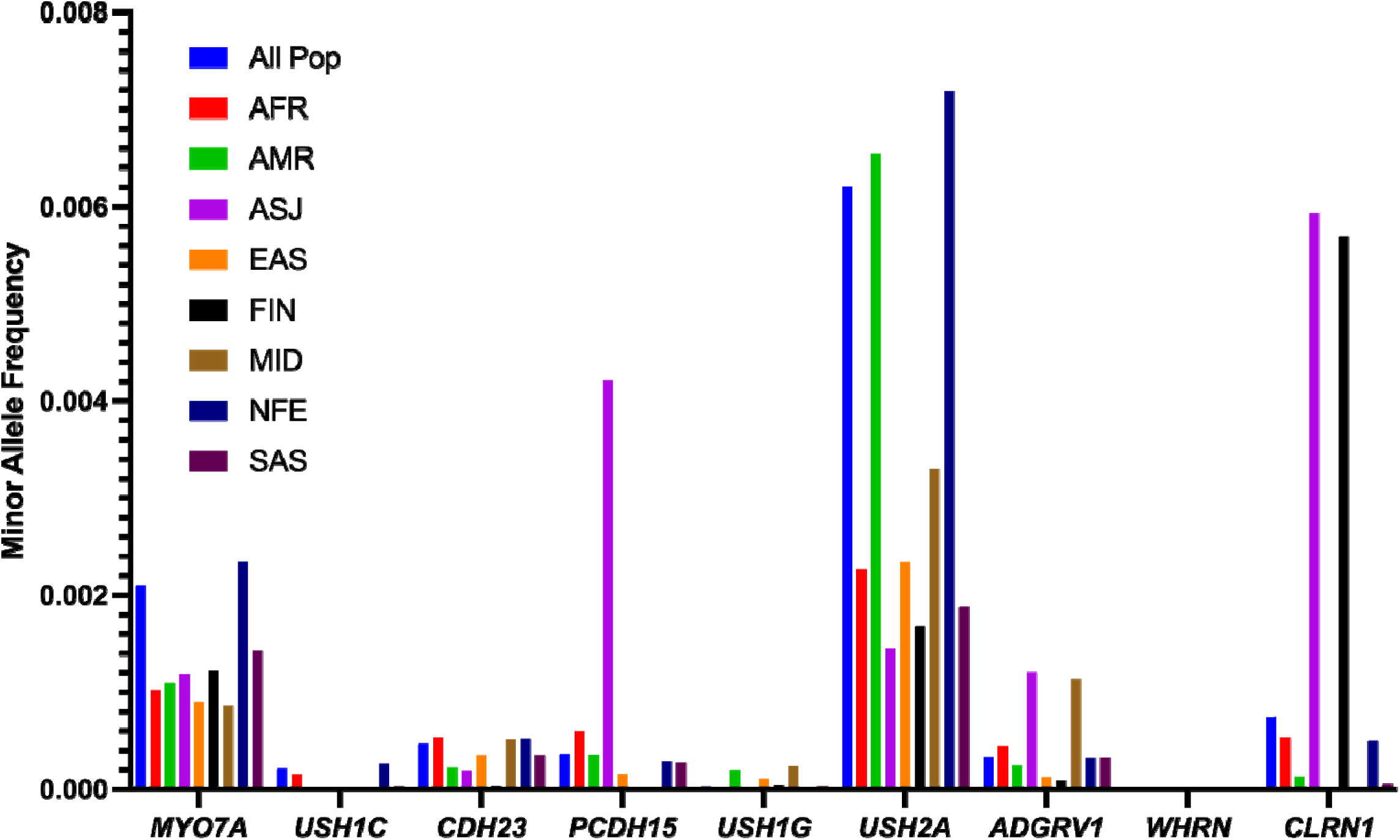
Sum of pathogenic and likely pathogenic Usher syndrome variants by genetic ancestry. Bars indicate the sum total of all minor allele frequencies for pathogenic and likely pathogenic variants per gene per ancestry population. Each gene is shown with populations along the x-axis and minor allele frequency on the y-axis. All pop, all populations; AFR, African/African American; AMR, Admixed American; ASJ, Ashkenazi Jewish; EAS, East Asian; FIN, Finnish; MID, Middle Eastern; NFE, Non-Finnish European; SAS, South Asian.

**Table 1.** The top ten most prevalent pathogenic/likely pathogenic Usher syndrome variants. Listed are population minor allele frequencies, sorted in descending order by all population frequency. The population with the highest minor allele frequency is highlighted. All pop, all populations; AFR, African/African American; AMR, Admixed American; ASJ, Ashkenazi Jewish; EAS, East Asian; FIN, Finnish; MID, Middle Eastern; NFE, Non-Finnish European; SAS, South Asian.

| Gene | Nucleotide Variation | Protein Variation | Consequence | Variation dbSNP ID | All Pop | AFR | AMR | ASJ | EAS | FIN | MID | NFE | SAS |
| --- | --- | --- | --- | --- | --- | --- | --- | --- | --- | --- | --- | --- | --- |
| *USH2A* | c.2276G>T | p.Cys759Phe | Missense | rs80338902 | 0.0015 | 0.0003 | **0.0018** | 0.0000 | 0.0000 | 0.0000 | 0.0007 | **0.0018** | 0.0000 |
| *USH2A* | c.10073G>A | p.Cys3358Tyr | Missense | rs148660051 | 0.0011 | 0.0001 | 0.0001 | 0.0000 | 0.0000 | 0.0000 | 0.0000 | **0.0013** | 0.0000 |
| *USH2A* | c.2299del | p.Glu767fs | Frameshift | rs80338903 | 0.0010 | 0.0003 | **0.0017** | 0.0000 | 0.0000 | 0.0000 | 0.0000 | 0.0012 | 0.0000 |
| *MYO7A* | c.3476G>T | p.Gly1159Val | Missense | rs199897298 | 0.0004 | 0.0001 | 0.0000 | 0.0000 | 0.0000 | 0.0000 | 0.0000 | **0.0005** | 0.0000 |
| *CLRN1* | c.300T>G | p.Tyr100Ter | Nonsense | rs121908140 | 0.0002 | 0.0000 | 0.0000 | 0.0000 | 0.0000 | **0.0057** | 0.0000 | 0.0001 | 0.0000 |
| *CLRN1* | c.118T>G | p.Cys40Gly | Missense | rs121908143 | 0.0002 | 0.0000 | 0.0000 | 0.0000 | 0.0000 | 0.0000 | 0.0000 | **0.0002** | 0.0000 |
| *CDH23* | c.5237G>A | p.Arg1746Gln | Missense | rs111033270 | 0.0002 | **0.0003** | 0.0000 | 0.0000 | 0.0000 | 0.0000 | 0.0000 | 0.0002 | 0.0000 |
| *CLRN1* | c.144T>G | p.Asn48Lys | Missense | rs111033258 | 0.0001 | 0.0000 | 0.0000 | **0.0059** | 0.0000 | 0.0000 | 0.0000 | 0.0000 | 0.0000 |
| *PCDH15* | c.733C>T | p.Arg245Ter | Nonsense | rs111033260 | 0.0001 | 0.0000 | 0.0002 | **0.0042** | 0.0000 | 0.0000 | 0.0000 | 0.0000 | 0.0000 |
| *CLRN1* | c.148_149insTGTC | p.Ser50fs | Frameshift | rs762606406 | 0.0001 | 0.0000 | 0.0000 | 0.0000 | 0.0000 | 0.0000 | 0.0000 | **0.0002** | 0.0000 |

**Table 2.**
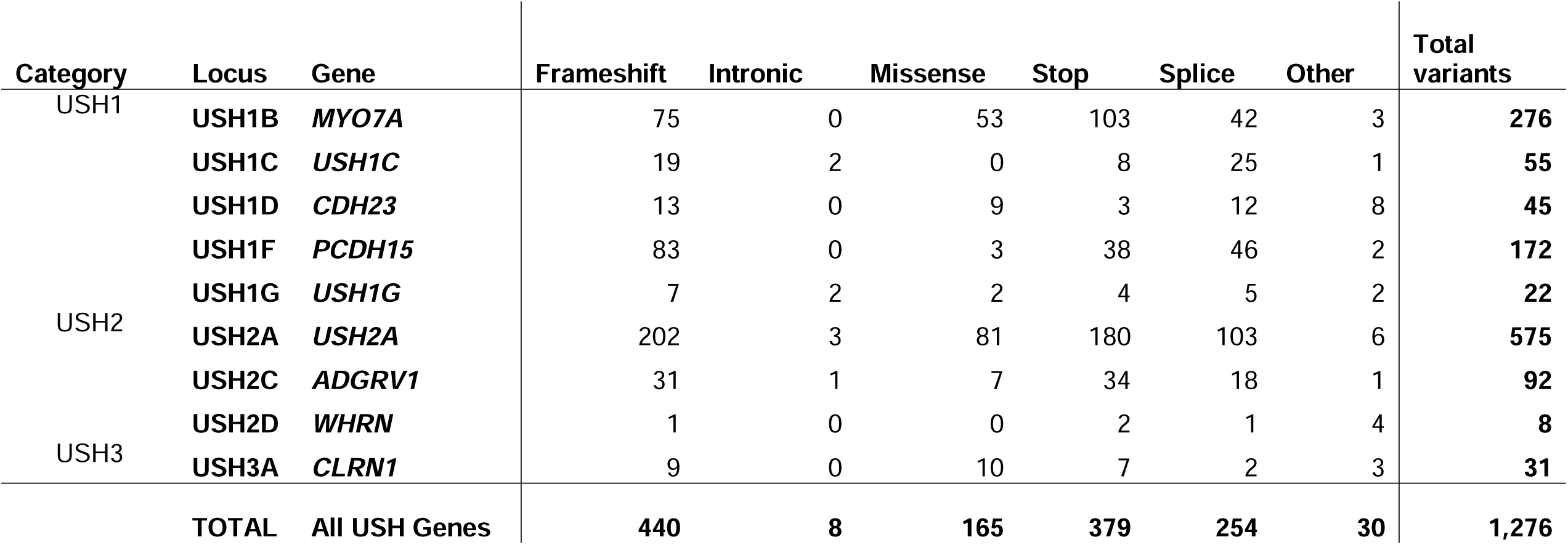
Pathogenic/Likely Pathogenic Usher syndrome variants by gene. Variants derived from ClinVar and included are only variants with ‘Usher syndrome’ included as the phenotype (ClinVar_USH, see methods).

### Prevalence of Usher Syndrome

We estimated prevalence of Usher syndrome using MAFs from 807,162 individuals (population control), 3,888 children without hearing loss (unaffected controls), as well as 637 children with hearing loss (affected population) using two statistical methods (Hardy-Weinberg and adjusted prevalence, see Methods). Given that the greatest variance in MAF total would arise from the number of annotated variants, we used five different databases of P/LP Usher variants which varied in the number of P/LP from 982 total (ClinVar_USH) to 5,860 (gnomAD_LoF) (see methods and **Supp Table 1**).

The population control database showed a carrier frequency of 0.010 for all Usher P/LP variants from ClinVar. P/LP carrier frequency for Usher type 1, 2, and 3 was 0.003, 0.007, and 0.001, respectively. The unaffected control database showed very similar figures: 0.010 overall Usher carrier frequency, 0.004 USH1, 0.006 USH2, and 0.0002 for USH3. The prevalence of Usher syndrome was 2.4% (15 of 637) of the children with hearing loss who underwent genetic testing (affected population), with seven diagnoses of USH1, seven of USH2, and one diagnosis of USH3.

The calculated prevalence of Usher syndrome in the general population ranged from 1 in 15,000 to 1 in 94,000 (**Supp Table 2**). The mean calculated prevalence across databases used for calculation was 1 in ∼29,000. For Usher syndrome type 1, 2, and 3, the mean calculated prevalence was 1 in ∼450,000, 1 in 50,000, and 1 in 8,000,000, respectively (**Supp Table 2**).

When extrapolated to the population of the United States, our results show that 30,405 individuals (range 3,585-88,614) are affected by Usher syndrome (**Figure 3** and **Supp Table 3**). When divided by subcategory, Usher syndrome type 1, 2, and 3, the calculated mean number affected (range) in the United States are 2,805 (278-5,955), 29,121 (3,260-82,488), and 66 (30-171), respectively.

**Figure 3.**
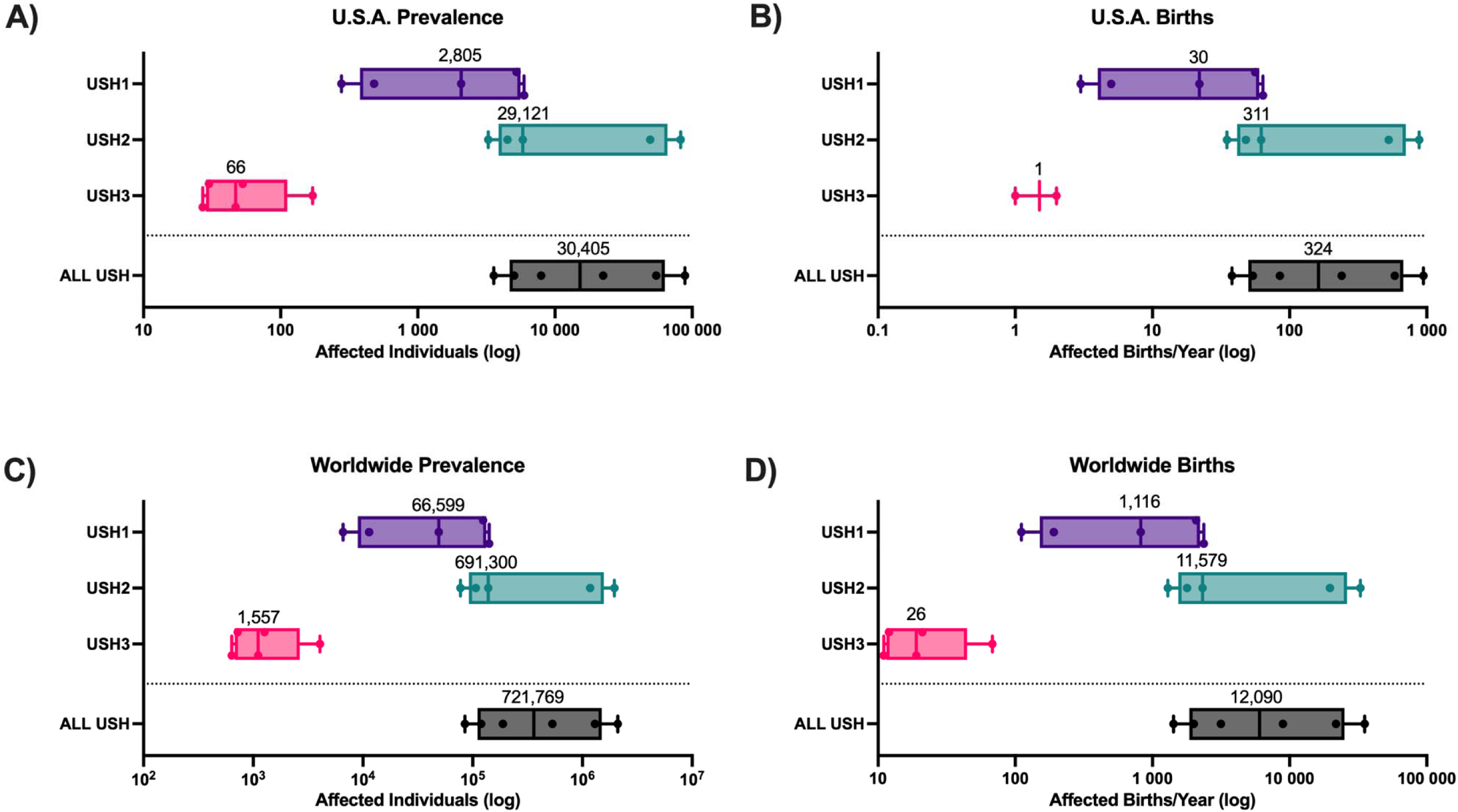
Prevalence and estimated births per year of Usher syndrome in the U.S.A. (A, B), and worldwide (C, D). Included are individual data points for each prevalence calculation (n = 6 calculations for ALL USH and n = 5 calculations for each USH subtype). Mean of calculations is printed above.

This indicates that the calculated mean for birth prevalence for Usher syndrome in the United States is 324 births/year (range 38-946). Worldwide, we calculated a disease prevalence of 721,769 (range 85,107-2,103,592) and a birth rate of 12,090 births/year (range 1,425-35,235) affected by Usher syndrome (**Figure 3** and **Supp Tables 4-6**).

### Pathogenic Variations Causing Usher Syndrome

The type of P/LP variants causing Usher syndrome varies by gene (**Figure 4**). Of note, as shown in **Figure 4B**, the number of reported P/LP variants does not reflect the actual contribution of these variants to disease at the population level. For example, ∼20% of reported P/LP variants in *PCDH15* are stop (nonsense) variants, however, these variants contribute to more than 50% of the prevalence of Usher syndrome caused by *PCDH15*. Similarly, while 20% of the reported P/LP variants in *MYO7A* are missense variant, they occur in over 60% of the USH1B population. Additionally, the frequency of intronic mutations associated with *USH1C* is the highest of all USH genes suggesting the presence of important regulatory motifs in the non-coding region of this gene.

**Figure 4.**
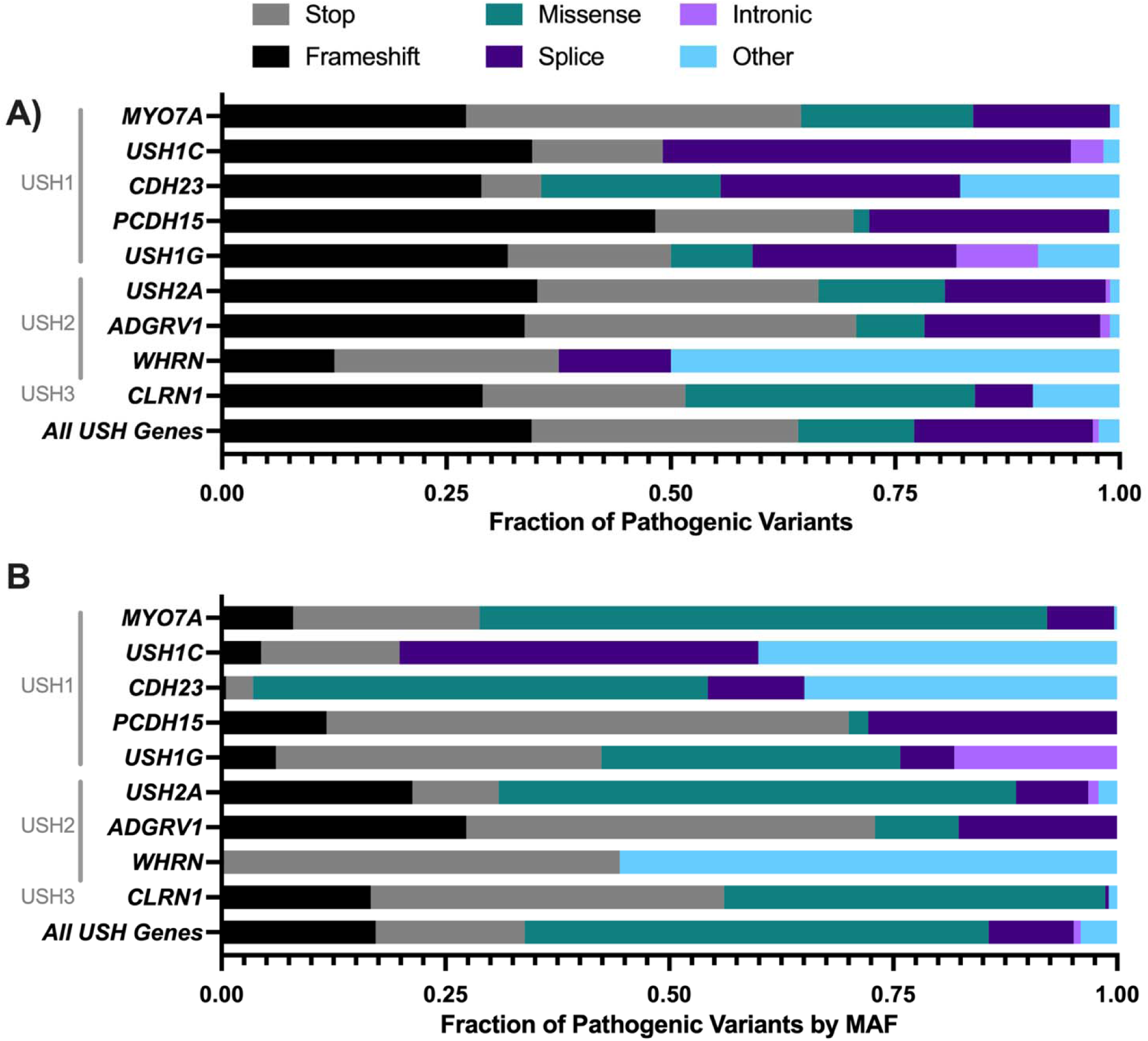
Types of Pathogenic Variations in Usher syndrome vary by gene (A) and contribution varies by Minor Allele Frequency (MAF) (B). Variants derived from ClinVar and included are only variants with ‘Usher syndrome’ included as the phenotype (ClinVar_USH, see methods).

### USH2A as a focus for alternatives to gene replacement therapy

*USH2A* is the most common genetic cause of Usher syndrome by MAF, carrier frequency, and number of reported pathogenic variants. The P/LP variants are distributed across 72 exons; however, variants in exon 13 contribute most significantly to the disease at the population level with a sum total minor allele frequency of 0.0025 (**Figure 5**).

**Figure 5.**
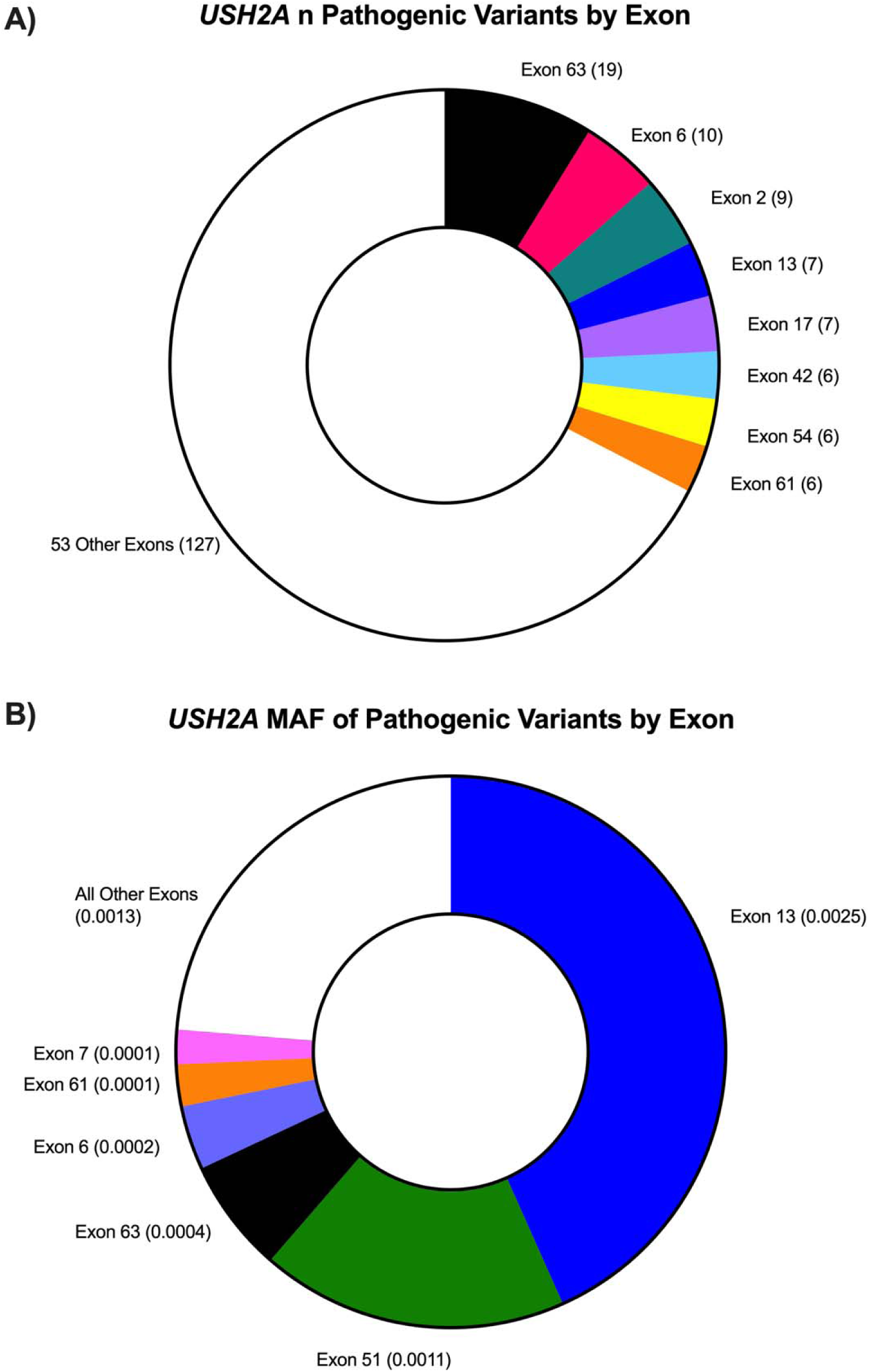
Pathogenic/Likely Pathogenic variations in *USH2A* by exon (A) and contribution varies by Minor Allele Frequency (MAF) (B). Variants derived from ClinVar and included are only variants with ‘Usher syndrome’ included as the phenotype (ClinVar_USH, see methods).

Although *USH2A* is the most common cause of Usher syndrome, it’s size precludes gene replacement therapy with currently available vectors. We characterized the 72 exons of *USH2A* based on whether they are in-frame, and thus possibly targetable by ASO-mediated exon skipping strategies (**Figure 6**). Twenty-six of 72 *USH2A* exons are in-frame, containing 152 (26%) of known P/LP variations. However, together these 152 variants make up a population-level MAF of 0.003, or 48% of the total P/LP variation contribution of all *USH2A* variants. Therefore, for *USH2A*, ASO strategies could be highly advantageous as a method for treatment that does not rely on gene-replacement. By this estimate, approximately 73% of individuals with *USH2A*-mediated Usher syndrome stand to benefit from ASO therapy, as only one of two mutated alleles must harbor a P/LP variant in an in-frame exon to be amendable.

**Figure 6.**
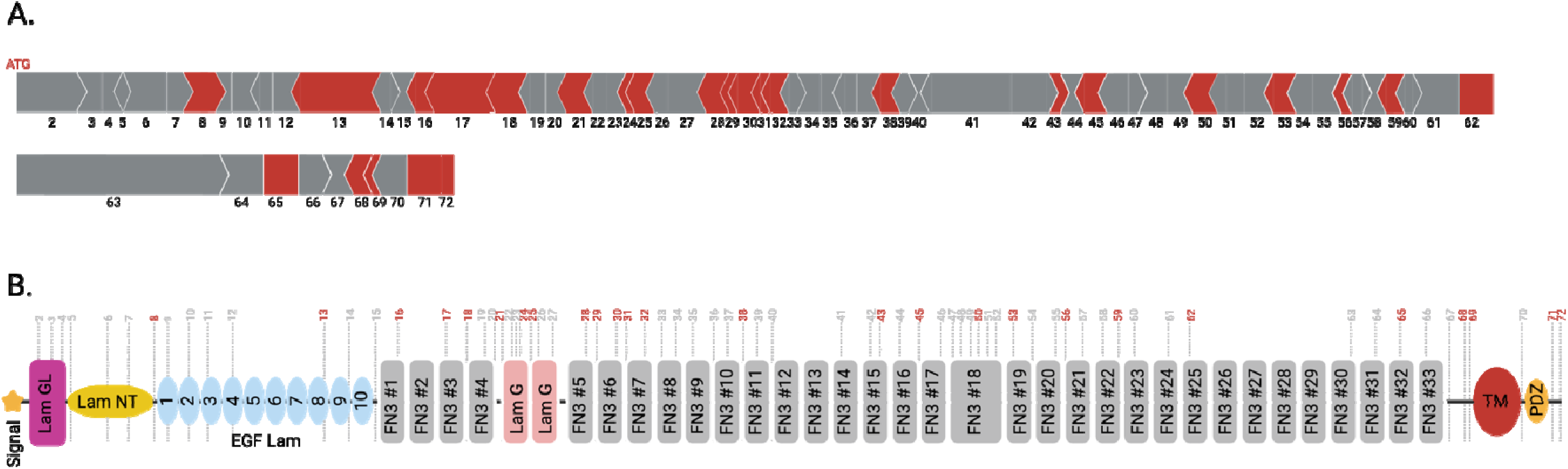
Schematic representation of the USH2A gene and protein, highlighting exons that can be targeted by antisense oligonucleotide (ASO)-mediated exon skipping strategies. (A) Schematic representation of the 72 exons (15,609 nucleotides) of the USH2A gene. In-frame exons (red) are potential targets for ASO-mediated exon skipping. Out-of-frame exons are indicated in grey. (B) Schematic representation of the USH2A protein (5,202 amino acids) based on SMART database (smart.embl-heidelberg.de). The upper numbers represent exons. The dotted lines depict the boundary of each exon that encode portions of the protein. Exons shown in red are in-frame and represent those that could be targeted by ASO-mediated exon skipping strategies, while exons in grey are out-of-frame.

## DISCUSSION

Usher syndrome is characterized by allelic, genetic, and phenotypic variability, making estimating the genetic contribution to disease and population-level prevalence estimates difficult. Prior estimates of prevalence were based either on small numbers of affected individuals (<50)^4,5^ which were then extrapolated to population level or analysis of patients presenting with isolated deafness^7^. Accurate data on the prevalence of Usher syndrome, including subtypes, are needed in order to guide molecular and genetic therapy development.

In this study, we provide the most accurate overall prevalence estimates for Usher syndrome based on P/LP variants derived from three clinical databases and evaluation of three populations: general population (gnomAD), unaffected population (3,888 children without hearing loss), and a large group of children presenting with isolated hearing loss (n=637). We found a carrier frequency of P/LP variants of 1% in both control populations. We calculate that the prevalence of Usher syndrome is 1 in ∼29,000. For Usher syndrome types 1, 2, and 3, the calculated prevalence was 1 in ∼450,000, 1 in 50,000, and 1 in 8,000,000, respectively.

The primary weakness of this study is reliance on reported P/LP Usher syndrome variants in public databases to make our calculations. We found wide variability in reported P/LP variants by database which would lead to vastly different calculations of prevalence, as a variant could only be annotated with a MAF if it is included in a database. We noted that the ClinVar_USH database of P/LP was the most stringent, with the fewest number of total P/LP Usher variants (n = 982) while the gnomAD_LoF was the least stringent with the number of variants (n = 5,860). When using these variants for calculations of prevalence, there was a wide range in sum total MAF (range 0.010 to 0.046). To begin to address this weakness, we included all five of these variant databases as well as the calculated adjusted prevalence, as previously described^4^, and derived the mean to provide the most accurate estimate of prevalence (**Figure 3**).

We noted significant variability on genetic contribution of Usher syndrome based on genetic ancestry (**Figure 1 & 2**). It is critical to take into account ancestry when evaluating the impact of genetic disorders on a population. For instance, P/LP variants in *PCHD15* are a major contributor to Usher syndrome in the ASJ population whereas *USH2A* is more common in NFE and AMR populations (**Figure 2**). If only the average of all populations is evaluated, the impact of a disorder like Usher syndrome in specific communities is not taken into account.

As discussed, due to the size of Usher syndrome genes, several are not amenable to gene replacement strategies^2,3,17^. And importantly, there is wide variability for genetic mechanism of disease for each gene. Here, we further characterize the wide distribution to genetic contribution to disease based on population MAF. By evaluating possible therapies in the context of MAF, we can better select impactful targets. Following our analysis, *USH2A* remains the most commonly affected USH gene in the United States and also Worldwide (**Figure 2 & 3**) with variants in exon 13 being the most frequent contributors (**Figure 5** and ^18,19^). *USH2A* is a very large gene with 72 exons and a coding sequence of 15,606bp which encodes usherin, a 5202 amino acid and 575Kda protein. In this case, neither gene replacement therapy or generation of a “mini” protein such as what has been developed and described for *PCDH15*^20^ can be envisaged. Instead, alternative methods must be explored such as the use of ASOs, designed to target a specific gene region to restore translation, in some cases skipping over entire exons. This approach has been successfully developed to target the most common mutation identified in exon 13^18^ and is currently undergoing clinical trial (clinicaltrials.gov). Single exon skipping should be considered for exons that are in frame (**Figure 6**). For other affected exons, multiple exon skips may be considered, bearing in mind potential for a deleterious impact of a larger deletion at the protein level. While in frame translation can be restored with an exon-skipping approach, the functionality of the altered protein remains to be demonstrated in most cases. Predictions can be made using deep learning protein modelling software and analysis of known functional and interacting regions of the proteins. However, validation of function in animal models remains the most reliable approach.

Prioritizing which variants to target will depend heavily on prevalence. As such, this report provides important insights into most comprehensive estimates of disease prevalence of Usher syndrome to date, highlighting the variability of Usher syndrome within different populations. Additionally, this report identifies key targets for molecular and gene therapy and will help guide future clinical work targeting specific variants in Usher syndrome genes.

## Data Accessibility Statement

Most data used in this analysis is available in publicly accessible databases or can be found in the work itself and supporting documents. Additional data related to this work may be requested from the authors.

## Acknowledgements

This work was supported by NIDCD K08 DC019716, the Boston Children’s Rare Disease Cohort Initiative, and the Boston Children’s Translational Research Program to AES. SAM and GSG receive support from the Rosamund Stone Zander Translational Neuroscience Center and the Manton Center at Boston Children’s hospital as well as from the Usher Syndrome Society and the Barber Gene Therapy Research Fund.

